# Efficacy of Corticosteroids in COVID-19 Patients: A Systematic Review and Meta-Analysis

**DOI:** 10.1101/2020.08.13.20174201

**Authors:** Haytham Tlayjeh, Olaa H. Mhish, Mushira A. Enani, Alya Alruwaili, Rana Tleyjeh, Lukman Thalib, Leslie Hassett, Yaseen M. Arabi, Tarek Kashour, Imad M. Tleyjeh

## Abstract

**Background:** To systematically review the literature about the effect of systemic corticosteroid therapy (CST) on outcomes of COVID-19 patients.

**Methods:** We searched Medline, Embase, EBM Reviews, Scopus, Web of Science, and preprints up to July 20, 2020. We included observational studies and randomized controlled trials (RCT) that assessed COVID-19 patients treated with CST. We pooled adjusted effect estimates of mortality and other outcomes using a random effect model, among studies at low or moderate risk for bias. We assessed the certainty of evidence for each outcome using the GRADE approach.

**Results:** Out of 1067 citations screened for eligibility, one RCT and 19 cohort studies were included (16,977 hospitalized patients). Ten studies (1 RCT and 9 cohorts) with 10,278 patients examined the effect of CST on short term mortality. The pooled adjusted RR was 0.92 (95% CI 0.69-1.22, I^2^=81.94 %). This effect was observed across all stages of disease severity. Four cohort studies examined the effect of CST on composite outcome of death, ICU admission and mechanical ventilation need. The pooled adjusted RR was 0.41(0.23-0.73, I^2^=78.69%). Six cohort studies examined the effect of CST on delayed viral clearance. The pooled adjusted RR was 1.47(95% CI 1.11-1.93, I^2^=43.38%).

**Conclusion:** Heterogeneous and low certainty cumulative evidence suggests that CST lacks efficacy in reducing short-term mortality while possibly delaying viral clearance in patients hospitalized with COVID-19. Because of the discordant results between the single RCT and observational studies, more research should continue to identify the clinical and biochemical characteristics of patients’ population that could benefit from CST.

## Introduction

The coronavirus disease 2019 (COVID-19) caused by the severe acute respiratory syndrome coronavirus 2 (SARS-CoV-2) continues to be a major global challenge with the paucity of proven effective therapies. In the majority of SARS-CoV-2 infected individuals, the disease is mild. In a proportion of patients, the immune response becomes dysregulated leading to severe acute lung injury manifesting as adult respiratory distress syndrome (ARDS) and can also lead to multiorgan injury and failure [1, 2].

The hallmark of pulmonary pathology in COVID-19 disease is diffuse alveolar damage, often associated with thickening of alveolar walls with infiltration by inflammatory cells dominated by macrophages and mononuclear cells [3]. It has been also observed that COVID-19 patients develop significant pulmonary vascular endothelial cell injury and endothelialitis, which is associated with intravascular thrombosis and microangiopathy [4, 5].

COVID-19 disease is commonly associated with several elevated inflammatory biomarkers, cytokines and chemokines reaching very high levels in the severe form. Among these are, C-reactive protein (CRP), ferritin, tumor necrosis factor-alpha (TNF), interleukins (IL-1, IL-2, IL-6, IL-8, and IL-10), Interferon-gamma, monocyte chemoattractant protein-1 (MCP-1) and granulocyte-macrophage colony stimulating factor (GM-CSF). Moreover, lymphocytopenia and neutrophilia are very common with a significant reduction in the CD8+ T cells, CD4+ T cells, and natural killer (NK) cell populations [1, 2].

The mortality among hospitalized patients ranges between 15% and 20%; however, it exceeds 40% in patients requiring intensive care [3]. Because of high in-hospital mortality, evidence of intense inflammation associated with COVID-19 disease and paucity of specific effective therapy, clinicians were forced to explore potential therapeutic approaches that target inflammation with the rationale to mitigate acute inflammation, reduce tissue injury and improve outcomes. Among the drugs that received early attention were corticosteroids because of their well-known broad-spectrum anti-inflammatory and immunomodulatory effects.

Corticosteroids therapy (CST) has been used extensively in acute respiratory conditions, which share similar pathological features with COVID-19 disease like SARS-CoV, MERS-CoV and H1N1 influenza, as well as in community acquired pneumonia (CAP), and ARDS. However, their effectiveness in reducing mortality and improving other outcomes in these conditions remain controversial [6-9]. Recent attempts to interpret these data led to more controversy regarding the therapeutic potential of CST in severe COVID-19 disease where some authors support and others recommend against their use in this disease [10, 11].

Early experience with CST in severe COVID-19 disease from a small observational study showed promising results in terms of improving survival [12], but subsequent observational studies and limited data from a single randomized clinical trial revealed mixed results. Since the single large randomized trial (RECOVERY) observed that dexamethasone treatment improves survival in the subgroup of patients with severe and critical COVID-19, CST has been promoted worldwide by physicians and the media as the most important level-I evidence effective COVID-19 therapy. Although a randomized controlled trial (RCT) is considered the gold standard to test the efficacy of any intervention, observational studies help to improve the inference derived from a single RCT, by improving generalizability. Although, the RECOVERY trial was a pragmatic trial with wide eligibility criteria, it used an open label design and it enrolled only 15% of hospitalized patients in the UK during the study period and excluded 17% of eligible patients because of unavailability of dexamethasone or treating physician’s decision. Data from observational studies help examine the generalizability of findings. In fact, empirical studies have shown that pooled estimates from meta-analysis of observational studies yield similar estimates to those pooled from RCTs [13, 14]. Moreover, there remain several questions that need to be addressed such as the timing of CST initiation, the dosing and the duration of treatment and disease phenotypes that could affect the efficacy of CST. Finally, observational studies may examine outcomes that are not assessed in RCTs. Therefore, we sought to perform a systematic review of randomized and observational studies addressing the role of CST in the treatment of COVID-19 disease and explore potential sources for heterogeneity of treatment effect in COVID-19 patients.

## Methods

We followed Preferred Reporting Items for Systematic Reviews and Meta-analyses (PRISMA) guidelines for reporting systematic review [15].

### Inclusion and Exclusion Criteria

We included 1) RCTs or 2) cohort or case-control studies reporting on the adjusted effect estimates of the association between CST use in COVID-19 patients and one of the following a-priori outcomes: (1) in-hospital mortality, (2) mechanical ventilation, (3) ICU admission, (4) viral shedding and (5) composite outcomes if reported.

### Data Sources and Search Strategies

A comprehensive search of several databases from 2019 to July 20, 2020, limited to English language and excluding animal studies, was conducted. The databases included Ovid MEDLINE(R) and Epub Ahead of Print, In-Process & Other Non-Indexed Citations and Daily, Ovid Embase, Ovid Cochrane Central Register of Controlled Trials, Ovid Cochrane Database of Systematic Reviews, Web of Science, and Scopus.

The search strategy was designed and conducted by an experienced librarian with input from the study’s principal investigator. Controlled vocabulary supplemented with keywords was used to search for studies describing CST for the treatment of COVID-19. The actual strategy including search terms used and how they were combined is listed in the supplementary material (Supplement AI: Search Strategy). We also searched for unpublished manuscripts using the medRxiv services operated by Cold Spring Harbor Laboratory and Research Square preprints. In addition, we searched Google Scholar and the references of eligible studies and review articles.

### Data Extraction

Four reviewers independently identified eligible studies (ME, AA, OM, RT) and extracted the data into a pre-specified data collection form. A senior reviewer verified all data included in the analyses (IT).

### Quality Assessment

Four reviewers (ME, AA, OM, HT) independently assessed the risk of bias for each study using the (RoB 2) of the Cochrane risk-of-bias tool for randomized trials and the ROBINS-I (“Risk Of Bias In Non-randomized Studies of Interventions”) for observational studies [16]. We also assessed all included studies for risk of survivor bias (or immortal time bias). Survivor bias occurs because patients who live longer are more likely to receive treatment than those who die early [17]. We considered the following analytical approaches as acceptable tools to account for survivor bias [8, 17, 18]: (1) CST use as a time-dependent variable in the regression analysis, (2) landmark analysis, (3) structural nested accelerated failure time model, (4) marginal structure models, and (5) matched cohort analysis in which each treated patient is followed up from the treatment start time with a matched control with the same disease duration prior to this time point. Studies that excluded patients who experienced the outcome within 24 hours of admission were considered at moderate risk for survivor bias. Reviewers judged each criterion for risk of bias and resolved any disagreements with a senior reviewer (IT). Finally, we assessed the certainty of evidence for each of our outcomes using the GRADE (Grading of Recommendations Assessment, Development, and Evaluation) approach [19, 20]. This method evaluates the certainty of evidence by assessing the following domains: Limitations, indirectness, inconsistency, imprecision, and publication bias.

### Statistical analysis

We evaluated between-studies heterogeneity using the I^2^ statistic which estimates the variability percentage in effect estimates that is due to heterogeneity rather than to chance—the larger the I^2^, the greater the heterogeneity [21]. Due to substantial heterogeneity, we pooled the adjusted effect estimates of included studies using the DerSimonian-Laird random-effects model and constructed corresponding forest plots [22]. Prior to pooling, the ORs were converted to RRs using the method by Zhang and Yu [23].

We conducted a priori determined subgroup analyses to assess the impact of (1) COVID-19 disease severity (critical group: patients admitted to intensive care unit (ICU). Severe group: patients requiring respiratory support outside ICU. Non-severe group: patients who did not require any respiratory support), (2) study design (RCT vs. cohort studies), (3) CST doses (low dose: methylprednisolone <= 1mg/kg/d or equivalence. High dose: methylprednisolone >1mg/kg/d or equivalence), (4) adjustment for survivor bias, on the overall estimate of effect. We also conducted meta-regression using study level baseline characteristics of patients’ populations and constructed corresponding bubble plots [24]. Due to missing data, we were able to examine only a few variables. Finally, we constructed contour-enhanced funnel plots and performed an Egger precision-weighted linear regression test as a statistical test of funnel plot asymmetry and publication bias [25]. All analyses were conducted using Stata version 16 statistical software (StataCorp, College Station, Texas).

## Results

### Included studies

A total of 24 studies (1 RCT, 23 cohorts) [26-49] with 16977 patients, including single center and multicenter studies from different countries, were included in our systematic review. Figure 1 shows the result of our search strategy (PRISMA flow diagram). Table 1 illustrates the general characteristics of the included studies. All studies reported on patients hospitalized with COVID-19 with varying degrees of disease severity.

**Figure 1:**
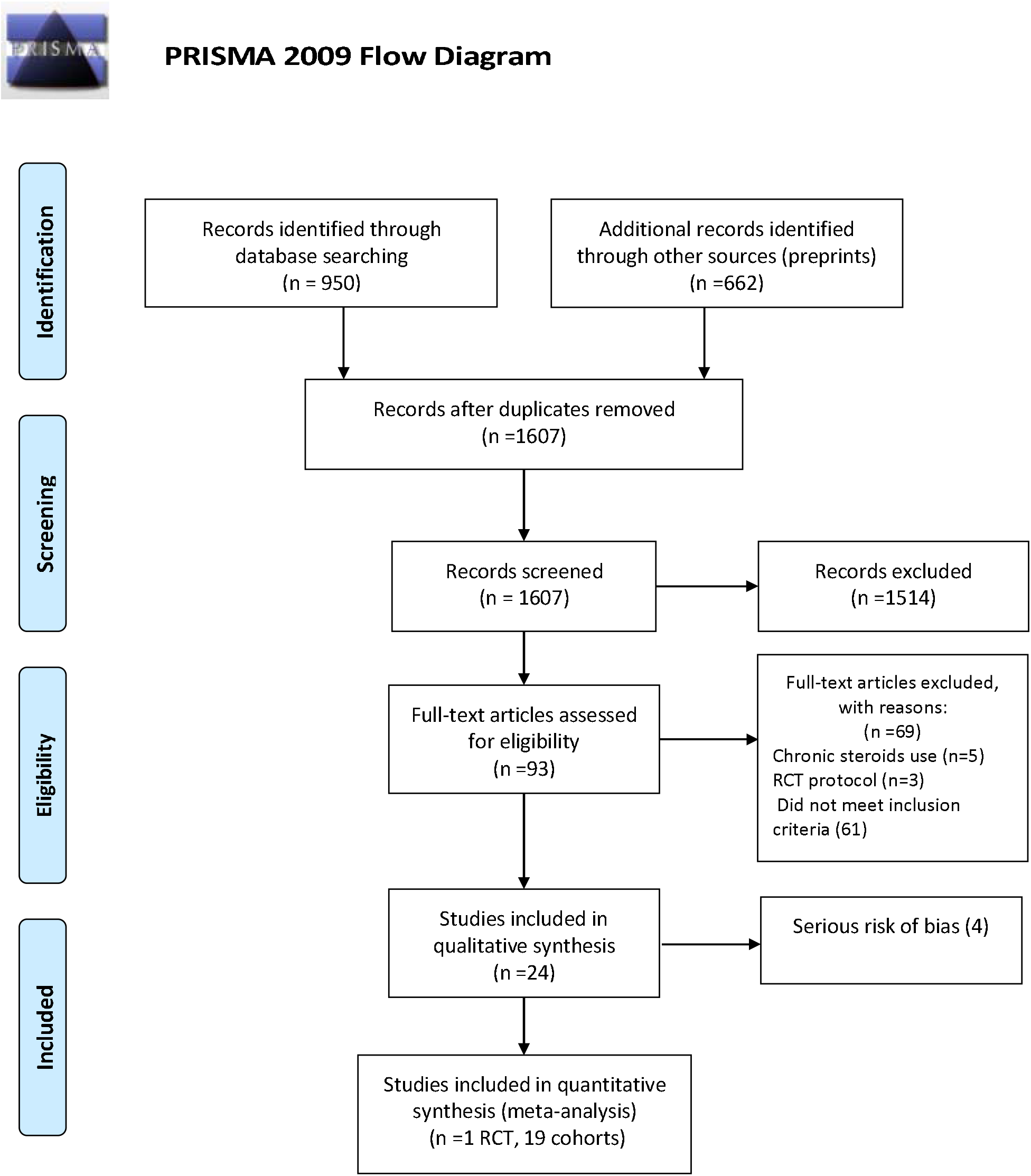
Flow Diagram of Included Studies

**Table 1.** Characteristics of Included Studies

| **Study/**<br>**Publication**<br>**year** | **Country** | **No. of**<br>**pts** | **Study type** | **Analytical**<br>**method** | **Cohort**<br>**selection** | **Exposure** | **Primary**<br>**outcome** | **Secondary**<br>**outcomes** | **Variables adjusted for** |
| --- | --- | --- | --- | --- | --- | --- | --- | --- | --- |
| **Albani et al,**<br>**2020** | Italy | 1403 | Retrospective<br>single center<br>cohort | multivariable<br>logistic<br>regression<br>with overlap<br>weight PS | Hospitalized<br>patients<br>with<br>moderate to<br>severe<br>COVID-19 | Corticosteroids,<br>dexamethasone<br>equivalent dose | In-hospital<br>Mortality | ICU admission | Age, sex, PaO<sub>2</sub>/FiO<sub>2</sub>,<br>lactate, CRP, platelets,<br>ICU admission,<br>antivirals |
| **Cao et al,**<br>**2020** | China | 58 | Retrospective<br>multicenter<br>cohort | Multivariable<br>Logistic<br>regression | Hospitalized<br>patients<br>with mild to<br>moderate<br>COVID-19 | Methyl<br>prednisolone 1-2<br>mg/kg per day | Progression<br>to severe<br>COVID-19 | NR | Age, gender, fever,<br>comorbidities, CRP,<br>lymphocytes count,<br>Time from onset of<br>illness to antiviral<br>treatment |
| **Chen et al,**<br>**2020** | China | 267 | Retrospective<br>single center<br>cohort | Multivariable<br>Cox-<br>regression<br>model | Hospitalized<br>patients<br>with COVID-19 | Corticosteroids | Prolonged<br>viral<br>shedding | NR | Age, time from<br>symptom onset to<br>admission, disease<br>severity, ICU, albumin,<br>oxygen therapy<br>diarrhea, antivirals and<br>antibiotic use |
| **Chroboczek**<br>**et al, 2020** | France | 70 | Retrospective<br>single center<br>cohort | Multivariate<br>logistic<br>regression<br>with PSM | Hospitalized<br>patients<br>with<br>moderate to<br>severe<br>COVID-19 | Corticosteroids | Orotracheal<br>intubation | NR | Age, sex, Charlson<br>index, BMI, time from<br>symptoms onset to<br>hospitalization, CRP,<br>HTN |
| **Corral-**<br>**Gudino et**<br>**al, 2020** | Spain | 85 | Partially<br>randomized<br>multi-center<br>open-label,<br>controlled<br>two-arm<br>group trial | Multivariate<br>adjusted risk<br>ratio | Hospitalized<br>patients<br>with<br>moderate to<br>severe<br>COVID-19 | Methyl<br>prednisolone<br>80 mg for 3 days,<br>then 40mg for 3<br>days | Composite<br>endpoint:<br>Mortality,<br>ICU<br>admission,<br>NIV<br>requirement | Laboratory<br>biomarkers,<br>individual<br>composite<br>endpoint | Age and baseline<br>respiratory status |
| **Crotty et al, 2020** | USA | 289 | Multicenter observational cohort | Multivariable Cox regression | Hospitalized patients with COVID-19 | corticosteroids | Respiratory bacterial co-infections | In-hospital mortality, ICU admission, mechanical ventilation, LOS | Age, ICU admission, MV, steroids, treatments, comorbidities, bacterial respiratory co-infections |
| **Fadel et al, 2020** | USA | 213 | Multicenter quasi-experiment | Multivariable logistic regression | Hospitalized patients with moderate to severe COVID-19 | Methyl prednisolone 0.5-1.0 mg/kg for 3-7 days | Composite endpoint: ICU admission, MV, in hospital mortality | Time to extubation, ARDS, shock, AKI, LOS | Age, gender, NEWS score |
| **Fernandez-Cruz et al, 2020** | Spain | 463 | Retrospective single center cohort | Multivariable logistic regression with PSM | Hospitalized patients with critical COVID-19 (ARDS and cytokine storm) | Methyl prednisolone 1 mg/kg/day or 0.5-1 gm/day | In-hospital mortality | NR | Age, comorbidities, ARDS severity, inflammatory markers |
| **Giacobbe et al, 2020** | Italy | 78 | Retrospective single center cohort | multivariable Cox regression | Hospitalized patients with critical COVID-19 | Methyl prednisolone 1 mg/kg daily | ICU-acquired BSI | Clinical characteristics and Predictors of BSI | Age, gender, comorbidities, hospital stay before ICU admission, SOFA score |
| **Horby et al, 2020** | UK | 6425 | Randomized, controlled, open-label trial | Cox regression | Hospitalized patients with COVID-19 | Dexamethasone 6mg daily for 10 days | 28-day mortality | Composite endpoint of invasive MV/ death; Hospital discharge | Age |
| **Li T.-Z. et al, 2020** | China | 66 | Retrospective single center cohort | Multivariable logistic regression | Hospitalized patients with COVID-19 | Methyl prednisolone | Prolonged viral shedding (>11 days) | NR | Age, disease severity, Fever, treatments, time from disease onset to admission |
| **Li X et al, 2020** | China | 548 | Ambispective single center | Multivariable Cox | Hospitalized patients | Prednisone equivalence 50 mg | In-hospital mortality | NR | Age, sex, LDH, WBC, complications, |
|  |  |  | cohort | proportional hazards regression | with moderate to severe COVID-19 |  |  |  | antivirals |
| **Liang et al, 2020** | China | 120/ 66 | Retrospective single center cohort | Case-control Propensity score matching | Hospitalized patients with critical COVID-19 | Methyl prednisolone | In-hospital Mortality | Duration of viral shedding, LOS | Age, gender, symptoms, inflammatory markers, comorbidities, LOS, MV |
| **Lu et al, 2020** | China | 244 | Retrospective single center cohort | Case-control Propensity score matching | Hospitalized patients with critical COVID-19 | corticosteroids | 28-day mortality | NR | Age, oxygenation, inflammatory markers |
| **Majmundar et al, 2020** | USA | 205 | Retrospective single center cohort | Multivariable Cox proportional hazards regression | Hospitalized moderate COVID-19 patients who developed acute respiratory failure outside ICU | corticosteroids | Composite endpoint: ICU admission, intubation, in-hospital mortality | Individual outcomes | Age, gender, oxygenation, comorbidities, inflammatory markers, treatments |
| **Narain et al, 2020** | USA | 2229 | Retrospective multicenter cohort | Multivariable Cox proportional hazards regression | Hospitalized critical COVID-19 with cytokine storm | corticosteroids | In-hospital mortality | NR | age, sex, race/ethnicity, insurance status, comorbidities, smoking, inflammatory markers, respiratory support |
| **Petrak et al, 2020** | USA | 145 | Ambispective multicenter cohort | Multivariable logistic regression with PSM | Hospitalized patients with moderate to severe COVID-19 | corticosteroids | Mechanical ventilation need | In-hospital mortality | age, sex, race, comorbidities, inflammatory markers |
| **Qi et al, 2020** | China | 147 | Retrospective single center | Multivariate logistic | Hospitalized patients | corticosteroids | Prolonged viral | NR | Symptoms, inflammatory markers, |
|  |  |  | cohort | regression | with COVID-19 |  | shedding (>17 days) |  | disease severity, treatments |
| **Salton et al, 2020** | Italy | 173 | Multicenter observational longitudinal cohort | Cox proportional hazards model | Hospitalized patients with severe COVID-19 | Methyl prednisolone protocol | Composite endpoint: 28-day mortality, ICU admission, orotracheal Intubation | MV-free days. Inflammatory markers changes | Age, sex, SOFA score, oxygenation, CRP level |
| **Shi et al, 2020** | China | 99 | Retrospective single center cohort | time-dependent Cox proportional hazard model | Hospitalized patients with COVID-19 | Corticosteroid 60 mg/d | Prolonged viral shedding | NR | Age, male sex, smoking, comorbidities, disease severity, Duration of illness, inflammatory markers |
| **Wang D et al, 2020** | China | 115 | Retrospective single center cohort | Multivariable logistic regression | Hospitalized patients with COVID-19 | Methyl prednisolone pulse or 1-3 mg/kg/d for 3-7 days | Composite endpoint: In-hospital mortality or ICU admission | NR | Age, male sex, comorbidities, inflammatory markers |
| **Wang K et al, 2020** | China | 68 | Prospective single center cohort | Cox proportional hazards model | Hospitalized patients with COVID-19 | Corticosteroids | Prolonged viral shedding | NR | Age, gender, comorbidities, duration of symptoms, respiratory support |
| **Wu J et al, 2020** | China | 1763 | Retrospective multicenter cohort | Multivariable Cox regression with time varying Propensity score match | Hospitalized patients with Severe & Critical Covid-19 | Corticosteroids | 28-d mortality | LOS, disease progression | Age, gender, inflammatory markers, oxygenation, comorbidities, smoking |
| **Xu K et al, 2020** | China | 113 | Retrospective multicenter | Multivariable logistic | Hospitalized patients | Methylprednisolone 0.5–1 mg/ kg/d | Prolonged viral | 21-d mortality | Age, gender, comorbidities, |
|  |  |  | cohort | regression | with COVID-19 |  | clearance (>15 days) |  | respiratory support, symptom duration |

Abbreviations: C-reactive protein, ICU: intensive care unit, NEWS: national early warning score, LOS: length of stay, MV: mechanical ventilation, SOFA: Sequential Organ Failure Assessment, NR: not reported.
\*Survivor Bias Adjustment: Wu and Shi studies performed time dependent cox regression. Fadel and Majmundar excluded patients with end points within $\leq 24$ hours. All others did not perform any survivor bias adjustment.

The quality of the observational studies was assessed using ROBINS-I tool (Figure 2B). Four studies were excluded from analysis because of serious risk of bias [27, 29, 36, 38]. Among included studies, immortal time bias was addressed in the analysis in only 3 (Table 2). The pooled RRs and corresponding ARRs and GRADE certainty of evidence are summarized in Table 2.

**Figure 2A:**
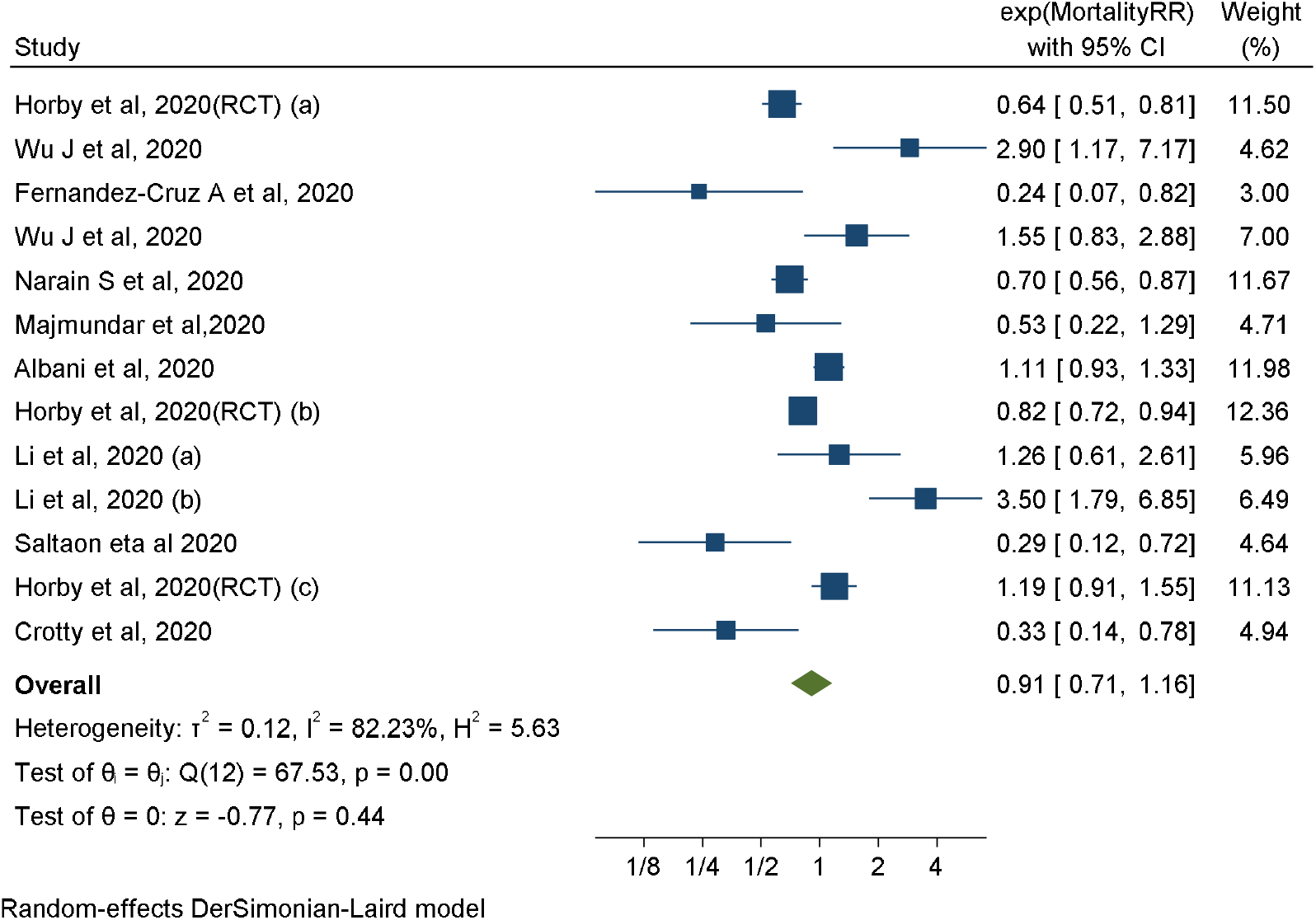
Association between corticosteroids use and short-term mortality in COVID-19 patients: (All cohorts and 1 RCT)

**Figure 2B:**
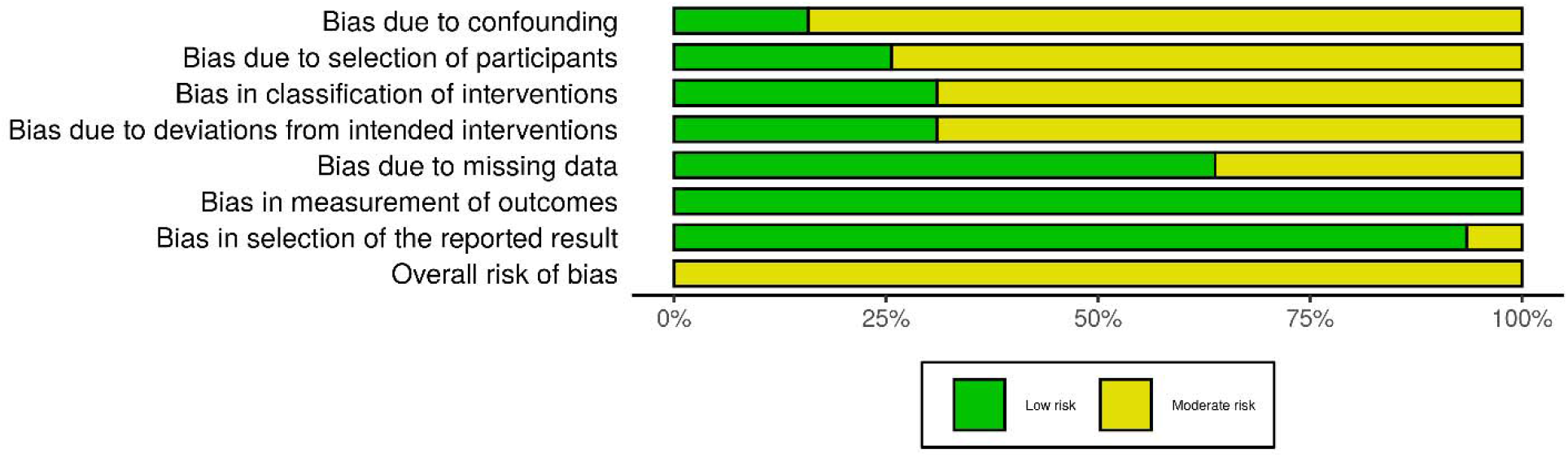
ROBINS-I quality assessment of included studies

**Table 2.** GRADE summary of findings: corticosteroids in patients with COVID-19, based on direct evidence from observational studies and one randomized controlled trial of patients with COVID-19

| Outcomes | Population | Relative effects (95%CI) | Baseline risk for control group | Absolute Risks Difference (95% CI) | Quality of evidence | Summary |
| --- | --- | --- | --- | --- | --- | --- |
| Mortality | COVID-19 with different disease severity | RR 0.92 (0.69 to 1.22) based on data from 10278 patients in 9 observational studies and 1 RCT | 19.9% | -1.6% (-6.2% to 4.4%) | Very low (serious inconsistency and imprecision) | We are very uncertain of the effect of corticosteroids on short term mortality in patients with COVID-19 |
| Mortality | Critical COVID-19 | RR 0.80 (0.26 to 2.46) based on data from 1719 patients in 2 observational studies and 1 RCT | 52.2% | -10.4% (-38.6% to 76.2%) | Very low (serious inconsistency) | We are very uncertain of the effect of corticosteroids on short term mortality in patients with critical COVID-19 |
| Mortality | Severe COVID-19 | RR 0.98 (0.73 to 1.30) from 9673 patients in 6 observational studies and 1 RCT | 15.8% | -0.3% (-4.3% to 4.8%) | Very low (serious inconsistency) | We are very uncertain of the effect of corticosteroids on short term mortality in patients with severe COVID-19 |
| Mortality | Non severe COVID-19 | RR 0.67 (0.19 to 2.34) based on data from 1824 patients in 1 observational study and 1 RCT | 14% | -4.6% (-11.4% to 18.8%) | Very low (serious inconsistency) | We are very uncertain of the effect of corticosteroids on short term mortality in patients with non-severe COVID-19 |
| Composite outcome of death/ICU/MV | Severe COVID-19 | RR 0.41 (0.23 to 0.73) based on data from 676 patients in 4 observational studies | 44.1% | -26% (-33.9% to 11.9%) | Very low (serious inconsistency) | We are very uncertain of the effect of corticosteroids on composite outcome of death/ICU admission/MV in patients with severe COVID-19 |
| Mechanical ventilation need | Severe COVID-19 | RR 0.74 (0.50 to 1.09) based on data from 5768 patients in 2 observational studies and 1 RCT | 9.3% | -2.4% (-4.7% to 0.8%) | Very low (serious inconsistency) | We are very uncertain of the effect of corticosteroids on mechanical ventilation need in patients with severe COVID-19 |
| Delayed Viral clearance | COVID-19 with different disease severity | RR 1.47 (1.11 to 1.93) based on data from 760 patients in 6 observational studies | 20.7% | 9.7% (2.3% to 19.2%) | Low | Corticosteroids may delay viral clearance in patients with COVID-19 |
| Acquired blood | COVID-19 with | HR 3.95 (1.20 to 13.03) | 45.5% | 45.4% | Very low | Corticosteroids probably do |
| stream infections | different disease severity | based on data from 78 patients in 1 observational study |  | (6.2% to 54.5%) | (serious inconsistency) | increase the risk of acquired blood stream infections in patients with COVID-19 |

### Mortality

Ten studies (1 RCT, 9 cohorts) [26, 31, 33, 35, 37, 39, 42, 46, 49] examined the effect of CST on short-term mortality in hospitalized patients with COVID-19. The pooled adjusted RR was 0.91 (95% CI 0.71-1.16, I^2^ = 82.23 %) indicating no significant association between CST and mortality (Figure 2A). There was significant heterogeneity between the studies. Contoured enhanced funnel plot showed no evidence of publication bias (Supplement AII: Figure1). On univariate meta-regression analysis, DM and male sex were associated with RR of mortality. The higher the prevalence of DM or male sex in included studies, the lower the reported RR for mortality (Supplement AII: Figure 2).

Three studies examined CST effect in patients with critical COVID-19 [26, 33, 46]. The pooled adjusted RR was 0.80 (95% CI 0.26-2.46, I^2^= 84.45%). Seven studies examined the effect in patients with severe COVID-19 [26, 35, 37, 39, 42, 46, 49]. The pooled adjusted RR was 0.98 (95% CI 0.73-1.3, I^2^=82.19%). Two studies examined the effect in patients with non-severe COVID-19 [26, 31]. The pooled adjusted RR was 0.67 (95% CI 0.19-2.34, I^2^=87.21%). There was no significant association between CST and short-term mortality across all the disease severity groups (Figure 3A). There was also no association between CST and mortality regardless of CST dose (pooled adjusted RR 0.95 (95% CI 0.74-1.22, I^2^ =78.59%) for low dose and RR 0.97 (95%CI 0.07-13.31, I^2^=92.98%) for high dose (Supplement AII: Figure 3).

**Figure 3A:**
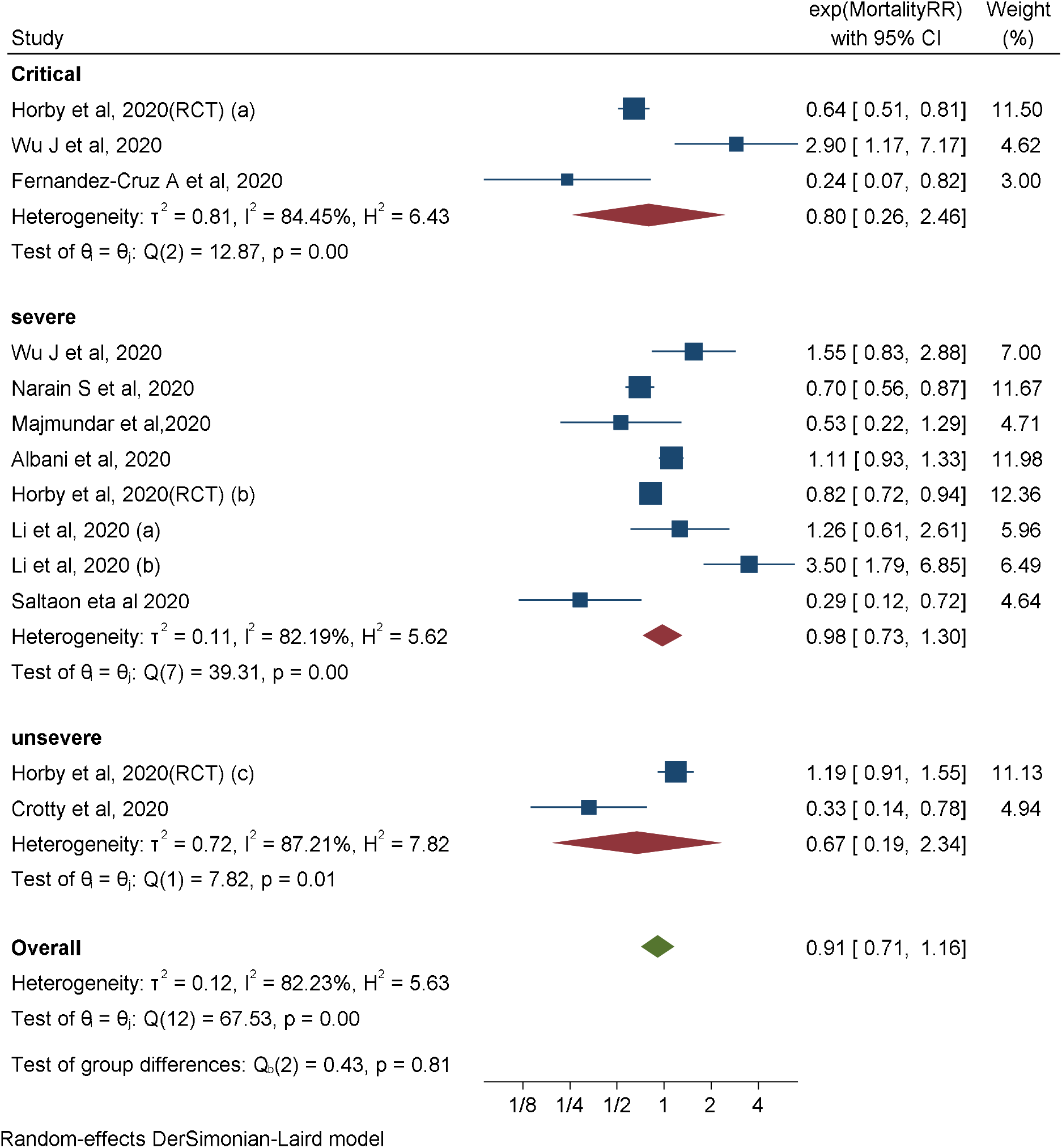
Association between corticosteroids use and short-term mortality in COVID-19 patients: By disease severity subgroups

**Figure 3B:**
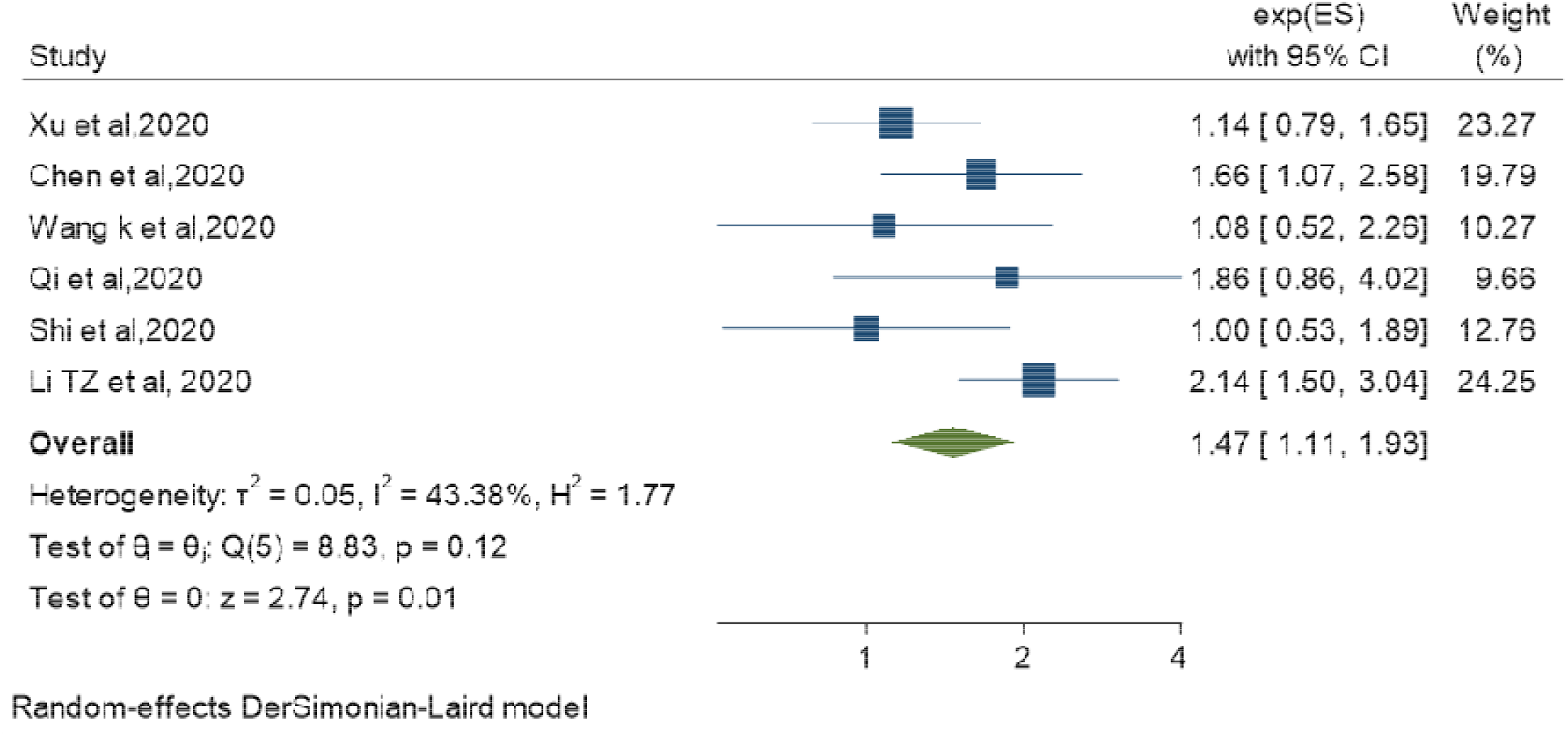
Association between corticosteroids use and delayed viral clearance in COVID-19 patients

Restricting the analysis to studies that adjusted for survivor bias showed no association between CST and mortality; pooled adjusted RR 1.03 (95% CI 0.75-1.42, I^2^ =82.49%) [26, 46] (Supplement AII: Figure 4).

### Composite outcome

Four cohort studies [30, 32, 37, 42] examined the effect of CST on composite outcome of death, ICU admission and mechanical ventilation need in patients with severe COVID-19. The pooled adjusted RR was 0.41(95% CI 0.23-0.73, I^2^=78.69%) (Supplement AII: Figure 5).

### Mechanical ventilation need

Three studies (1 RCT, 2 cohorts) [26, 37, 40] showed no association between treatment with CST and mechanical ventilation need. The pooled adjusted RR was 0.74 (95% CI 0.50-1.09, I^2^=74.15%) (Supplement AII: Figure 6).

### Viral clearance

Six cohort studies examined the effect of CST on viral clearance [28, 41, 43, 45, 47, 48]. Viral clearance is defined as two consecutive negative RT-PCR swabs separated by at least 24 hours. Delayed viral clearance was not uniformly defined but most studies defined it as persistent positive RT-PCR swabs for more than 11 to 17 days from first positive test. The six included patients with COVID-19 with variable degree of disease severity. CST was associated with delayed viral clearance; pooled adjusted RR was 1.47 (95% CI 1.11-1.93, I^2^=43.38%) (Figure 3B). The RECOVERY trial did not assess the effect of CST on viral clearance.

## Discussion

### Main findings

This systematic review and meta-analysis included 19 cohorts and 1 RCT, with low to moderate risk of bias, which addressed the association between CST and mortality, disease progression, and viral clearance in patients hospitalized with COVID-19 disease. Although the single pragmatic RECOVERY randomized trial showed that CST was associated with lower mortality in severe and critical COVID-19, we could not observe the same effect in our large meta-analysis of 16977 patients. We found with very low level of certainty, that CST did not reduce short-term mortality among COVID-19 patients. However, in a smaller number of studies (4 cohorts) that reported on composite outcomes, we found with very low level of certainty (due to heterogeneity) an association between CST and a decreased risk of the composite outcome (death, ICU admission, and mechanical ventilation need). These observations did not change when we restricted our analysis to studies that adjusted for survivor bias. We also found similar results across all stages of disease severity, critical, severe, and non-severe. All meta-analyses were limited by significant between-studies heterogeneity that could not be explained by disease severity, high vs. low dose of corticosteroids. On the other hand, we found that CST could prolong viral shedding, and in a single study that examined the risk of secondary infections, CST was associated with an increased risk of acquired bloodstream infections [34].

### Discordance between the RCT and observational studies

Although the cumulative evidence from our meta-analysis is discordant with the single RCT (RECOVERY), it should not be disregarded as inferior evidence in the presence of this RCT. First, the RECOVERY trial had an open label design which could make it susceptible to performance (co-intervention) bias that usually inflicts observational studies. Second, it was conducted in a single country and the baseline mortality for patients with mild and severe disease was higher relative to that reported from other countries which makes the results less generalizable to different patients’ populations. Third, although the RECOVERY trial due to its pragmatic design was an exceptional achievement during the pandemic, only 15% of hospitalized patients in UK were enrolled and 17% of eligible patients were excluded from enrollment in this RCT because dexamethasone was either not available, or it was believed to be indicated or contraindicated by the treating physicians. Fourth, despite compelling evidence from RCTs, observational studies if appropriately designed and analyzed provide important evidence from real-life data. Observational studies and meta-analyses of these studies may offer higher external validity than a single RCT owing to their potentially large size and the ability to include a patient sample that is representative of the average patient population. Fourth, there are additional important differences between RCTs and observational studies, such as standardized patients care with protocols and exclusion of certain patients’ groups in RCTs [50].

### Comparison to other pneumonia and lung injury syndromes

Our findings of lack of efficacy of CST in reducing mortality among COVID-19 patients is concordant with previous observations in other coronavirus infections associated acute lung injury, namely SARS-CoV and MERS-CoV. In a systematic review of 29 studies of CST in SARS-CoV infection, the results of 25 studies were inconclusive while four studies showed possible harm [7]. Similarly, in a recent systematic review that included two large cohorts with 6129 SARS-CoV patients, CST did not show a significant reduction in mortality (HR 0.83, 95% CI 0.41-1.66) [9]. Lack of efficacy of CST in reducing mortality was also shown in MERS-CoV infection. In a multicenter study of 309, the crude mortality was higher in the corticosteroid treated group (74.2% vs. 57.6%, p=0.002) but the adjusted mortality was not different (adjusted OR 0.78, 95% CI 0.52-1.07, p=0.12) [8]. On the other hand, CST was associated with increased mortality among influenza A H1N1 patients. In a systematic review and meta-analysis of 10 observational studies involving 6548 patients, CST was associated with increased (risk ratio [RR] 1.75, 0.95% CI 1.30-2.36, p=0.0002) [51]. Conversely, a systematic review of RCTs showed that CST improved survival in patients admitted with severe community-acquired pneumonia [52]. The discrepant response to CST between bacterial and viral pneumonias may be due differences other than the study design such as immuno-inflammatory phenotypic differences between viral and bacterial pneumonia. Moreover, effective antimicrobial therapy in the case of bacterial pneumonia may have influenced the findings.

### Different disease phenotypes

Many studies have described different immune phenotypes among COVID-19 patients particularly after day 10 of the infection. Patients may exhibit different immune trajectories despite similar clinical severity at initial presentation. It was observed that one patient population exhibits low expression of proinflammatory cytokines and enrichment in tissue repair genes and in another COVID-19 group, patients showed persistent elevation of proinflammatory cytokines and progressed to develop cytokine release syndrome (CRS) [53].

In addition, autopsy series from USA and Italy identified different pathological phenotypes in patients who died from COVID-19 disease. Some patients had extensive alveolar inflammation with diffuse alveolar damage, while others had extensive endothelial damage with intravascular platelet-fibrin thrombi leading to organ dysfunction without alveolar inflammation and injury [54, 55].

Gattinoni et al also described different non-uniform clinical phenotypes of COVID-19 infections with different disease characteristics that necessitate different treatment modalities [56]. Many critically ill patients who are very hypoxemic have extensive alveolar inflammation and typical ARDS while others have no evidence of alveolar disease on chest imaging.

In the RECOVERY trial, patients on respiratory support benefited most from CST; however, there was possible harm in patients who were not on oxygen. In a large observational cohort study that adjusted for immortal time bias, Wu et al found increased mortality with CST in severe and critical COVID-19 patients. This highlights the heterogenous response of patients with COVID-19 to different immunomodulators, especially corticosteroids. The effect of phenotypic differences on response to immunomodulating therapies has been seen in ARDS and sepsis patients treated with simvastatin and anakinra respectively. In a post hoc analysis of the HARP-2 trial, ARDS patients with hyperinflammatory sub-phenotype but not those with hypo inflammatory sub-phenotype responded favorably to simvastatin therapy [57]. Likewise, in a post hoc analysis of an RCT of sepsis patients treated with anakinra (IL-1b receptor antagonist), only patients with evidence of hepatic dysfunction and disseminated intravascular coagulation had significant reduction in 28-day mortality (HR 0.28, 95% CI 0.11-0.71, p=0.007) [58].

The patients in our meta-analysis were very heterogeneous and that could explain the lack of efficacy of CST in improving outcomes in general but could be helpful in certain phenotypes as shown in the subgroup analysis of the RECOVERY trial.

We could not explore all possible causes of between-studies heterogeneity with the meta-regression analysis such inflammatory markers levels (CRP, D-Dimer, IL-6) because of inadequate data reporting in included studies.

### Dose of corticosteroids

The dose of corticosteroids used in the RECOVERY trial was dexamethasone 6 mg per day (methylprednisolone equivalence 32 mg, <0.5mg/kg/d for an average 70 kg person). All the cohort studies included in our systematic review used higher doses of steroids. High dose corticosteroids are associated with more serious adverse effects like serious hyperglycemia, gastrointestinal bleeding, neuropsychiatric events, and superinfection [9]. However, looking at subgroups of studies that used <=1 mg/kg/d vs >1 mg/kg/d of methylprednisolone equivalence showed no effect on mortality regardless of the dose used. Consistent with our observation, Lansbury et al conducted a meta-analysis on the effect of adjunctive corticosteroid therapy in influenza and showed no clear association between corticosteroid dose and mortality [59]. On the other hand, in a systematic review of 4 RCTs and 5 cohort studies of ARDS patients, low dose CST was associated with improved mortality [60]. Li et al showed higher mortality with high dose corticosteroids in severe COVID-19, adjusted RR 3.5 (1.79-6.85). This highlights the importance of the dose of steroids used in COVID-19 infection to attenuate the inflammatory response, avoiding serious adverse effects that can negate any potential benefit.

### Timing of corticosteroids administration

SARS-CoV2 viral replication starts to decline after the first week of infection with the peak of interferon levels [61]. The COVID-19 immune signature starts to change in the second week with possible dysregulated immune system trajectory as described earlier.

In a priori subgroup analysis of the RECOVERY trial, the patients who benefited most from CST are patients who were commenced on CST more than 7 days from onset of symptoms which may correlate with the start of dysregulated immune system. We could not assess the effect of CST timing in our systematic review because of inadequate data. The timing of CST in the disease stage likely play a vital role in modulating the immune response and outcome.

### Prolonged Viral Shedding

Lucas et al found that nasopharyngeal viral load correlates positively with plasma levels of interferon and cytokines. Patients with severe disease did not show any decline in viral load over the course of their disease [53]. We observed a significant association between CST and prolonged viral RNA shedding, pooled RR 1.47 (1.11-1.93) which is consistent with previous data in other viral infections such as SARS-CoV, MERS-CoV and influenza [7, 8, 62]. Prolonged viral RNA shedding is often used as a surrogate for viral replication but the correlation between prolonged viral RNA shedding and infectivity and other clinical outcomes is unknown.

### Strengths and Limitations

Our meta-analysis has several strengths. Firstly, published, and unpublished studies were included, which reduces publication bias. We also employed rigorous methodologies. We excluded studies that were prone to significant confounding because they did not report adjusted odds or hazard ratios. We also examined mortality and other clinical outcomes separately and performed sensitivity analyses to explore sources of between studies heterogeneity. However, our study has several limitations; all our included studies except one were observational studies which are prone to different biases; including confounding by indication, survivor (immortal time) bias and residual confounding. Our group and others have shown that survivor bias, which occurs because patients who live longer are more likely to receive treatment than those who die early, could change associations from benefit to harm [8, 17, 63]. Only one observational study has adjusted for survivor bias [46] and in this single study, CST was associated with a higher mortality in both severe and critical subgroups. Moreover, as with all observational studies, residual confounding could inflict any observed association [64] even with appropriate adjustment or propensity score matching. Nevertheless, the direction of these different biases is supposed to be in favor of corticosteroids efficacy, which was not observed in our analysis.

### Conclusions

We found in this systematic review and meta-analysis that heterogeneous and low certainty cumulative evidence suggests that CST lacks efficacy in reducing short-term mortality while possibly delaying viral clearance in patients hospitalized with COVID-19. Because of the discordant results between the single RCT and observational studies, more research should continue to identify the clinical and biochemical characteristics of patients’ population that could benefit from CST and the optimal dose and duration of treatment.

## Data Availability

All data and materials generated during the current study are available from the corresponding author on reasonable request

## Authors’ contribution

HT and IT designed the study. LH performed literature search. ME, AA, OM, LH and RT performed literature screening, study selection and data extraction. HT, IT, ME, AA, and OM assessed the risk of bias. LT and IT conducted the statistical analyses. HT, IT, YA, and TK led the writing of the manuscript. IT and YA performed data interpretation. All authors revised the manuscript for important intellectual content.

## Funding

None

## Conflicts of interest

IT reports personal fees from UpToDate, outside the submitted work.

YA reports that he is principal investigator on a clinical trial of lopinavir–ritonavir and interferon for Middle East respiratory syndrome (MERS) and that he was a non-paid consultant on therapeutics for MERS-coronavirus (CoV) for Gilead Sciences and SAB Biotherapeutics. He is a co-investigator on the Randomized, Embedded, Multi-factorial Adaptive Platform Trial for Community-Acquired Pneumonia (REMAP-CAP) and a board member of the International Severe Acute Respiratory and Emerging Infection Consortium (ISARIC).

The other authors declare no competing interests.

## Ethics approval

No ethics approval was obtained because data from previous published studies were compiled and analyzed.

## Consent to participate

Not applicable.

## Consent for publication

All authors approved the final version submitted for publication

## Notes

### Competing Interest Statement

The authors have declared no competing interest.

